# Comparison between the Smart Triage model and the Emergency Triage Assessment and Treatment (ETAT) guidelines in triaging children presenting to the emergency departments of two public hospitals in Kenya

**DOI:** 10.1101/2023.11.08.23298265

**Authors:** Stephen Kamau, Joyce Kigo, Paul Mwaniki, Dustin Dunsmuir, Yashodani Pillay, Cherri Zhang, Brian Nyamwaya, David Kimutai, Mary Ouma, Ismael Mohammed, Keziah Gachuhi, Mary Chege, Lydia Thuranira, J Mark Ansermino, Samuel Akech

## Abstract

Several triage systems have been developed, but little is known about their performance in low-resource settings. Evaluating and comparing novel triage systems to existing triage scales provides essential information about their added value, reliability, safety, and effectiveness before adoption. This prospective observational study included children aged < 15 years who presented to the emergency departments of two public hospitals in Kenya between February and December 2021. We compared the performance of Emergency Triage Assessment and Treatment (*ETAT*) guidelines and Smart Triage (ST) models (*ST-only model*, *ST model with independent triggers*, and *recalibrated ST model with independent triggers*) in categorizing children into emergency, priority, and non-urgent triage categories. We visualized changes in classification of participants using Sankey diagrams. 5618 children were enrolled, and the majority (3113, 55.4%) were aged between one and five years of age. Overall admission and mortality rates were 7% and 0.9%, respectively. *ETAT* classified less children, 513 (9.2%), into the emergency category compared to 790 (14.1%), 1163 (20.8%) and 1161 (20.7%) by the *ST-only model*, *ST model with independent triggers* and *recalibrated model with independent triggers*, respectively. *ETAT* also classified more children, 3089 (55.1%), into the non-urgent triage category compared to 2442 (43.6%), 2097 (37.4%) and 2617 (46.7%) for the respective *ST models*. *ETAT* classified 191/395 (48.4%) of admitted patients as emergency compared to more than half by all the *ST models*. *ETAT* and the *ST-only model* classified 25/49 (51%) children who died as emergencies, while the *ST models with independent triggers* classified 39/49 (79.6%) children as emergencies. Smart Triage shows potential for identifying critically ill children in low-resource settings, particularly when combined with independent triggers. Additionally, it performs comparably to *ETAT*. Evaluation of Smart Triage in other contexts and comparison to other triage systems is required.

**Author summary:** Prioritizing children according to the level of severity of illness in the outpatient department is crucial to ensure very sick children are identified and receive life-saving treatment while those with less severe symptoms can safely wait in the queue. Appropriate triage prevents avoidable paediatric mortality. As new triage systems are developed, it is essential to evaluate their performance before being used by healthcare professionals to manage patients. In this study, we compared a newly developed triage algorithm, Smart Triage, to the World Health Organization’s Emergency Triage Assessment and Treatment (*ETAT*) guidelines. Here, we highlight how participants were categorised into emergency, priority, and non-urgent categories by both triage systems. We also assessed changes in triage categorization by comparing the Smart Triage model only (with and without site specific recalibration) and the model with independent emergency and priority triggers aligned with *ETAT*. Our study shows that Smart Triage had comparable performance to *ETAT,* and it can be used to triage children in resource-limited settings. Smart Triage can be integrated into a digital device allowing frontline healthcare workers to rapidly triage children presenting to the outpatient department and recognize very sick children faster, so that they can be treated in a timely manner.

## Introduction

Overcrowding is a major global problem in many emergency departments (ED) (1,2). This is compounded by lack of validated triage systems that can help health workers distinguish between low and high-priority patients, along with poor adherence to existing triage protocols (3). Triage is a vital component of effective and efficient emergency care for children presenting to health facilities with varying severity of illness. Triage categorizes patients according to the severity of their illness and designates a level of urgency that includes non-urgent, priority, and emergency cases that require immediate medical attention (4).

Rapid triage of critically ill patients can reduce the waiting time for children needing life-saving treatment by distinguishing them from non-urgent patients who can safely wait in the queue for assessment and treatment by health workers (5,6). However, in resource-limited settings, triage remains underused, especially in paediatric emergency care, owing to significant barriers such as inadequate staffing, the complexity of guidelines, social and organizational context, and lack of capacity (7–9). Consequently, the sickest children are not prioritized, resulting in delayed care and inefficient resource utilization.

Several paediatric triage systems have been developed and adopted in high and low-income settings to help frontline health workers improve triage accuracy (10–14). One of the most widely implemented triage systems is the World Health Organization’s (WHO) Emergency Triage Assessment and Treatment (*ETAT*) guidelines, which are recommended for use in resource-constrained settings (15,16). The *ETAT* system provides a systematic and objective approach to triaging children using clinical signs to identify emergency, priority, and non-urgent cases. However, the implementation of *ETAT* in clinical practice has faced a myriad of challenges, including a high turnover of trained staff, lengthy and high-intensity training, and limited resources to support implementation (17,18).

A new paediatric triage model, Smart Triage, was recently developed based on the data collected from a hospital in Uganda (19). This is a logistic regression model based on nine variables that were selected from over 100 demographic, vital sign measurements, symptoms, and socio-demographic variables collected upon arrival at the hospital. This model provides the probability of admission. The prediction model addresses some of the challenges associated with the implementation of *ETAT* by utilizing a limited number of predictors to identify critically ill children and can be used by frontline health workers with limited training and expertise. Moreover, the model can be integrated into digital health platforms to minimize the need for memorization of triage protocols and training of triage staff. Implementation of the model as a triage system should also incorporate a set of independent emergency and priority triggers, as in other triage systems, such as *ETAT*. These triggers include rare emergency conditions and ensure that children who would have been misclassified by the model are safely assigned to either emergency or priority categories.

It is imperative to evaluate the performance of new triage systems and, if possible, to compare them with existing systems before clinical adoption. Assessment of a triage system’s performance should be based on its ability to differentiate between low and high acuity patients as they present to the ED, thus minimizing misclassification of patients. Misclassification of low acuity patients into the high acuity category misuses scarce resources and increases the waiting time for more urgent patients. On the other hand, misclassification of high acuity patients into a low acuity category may result in delayed assessment, diagnosis, and treatment, and potentially poorer outcomes (4,20). Compared with existing paper-based guidelines, electronic triage systems provide valuable information to policymakers and health workers regarding the safety, reliability, and effectiveness of a triage system within a particular setting. Furthermore, potential research gaps and areas of improvement can be identified during the evaluation.

The purpose of this study was to compare the performance of *ETAT* to Smart Triage in classifying children into triage categories using prospectively collected paediatric data from two Kenyan public hospitals. This comparison can provide validation and performance trade-offs of the new triage model compared with the *ETAT* algorithm and inform its utility in low-resource settings.

## Materials and Methods

### Study design

This prospective observational study was conducted at the emergency department (ED) of Mbagathi County Hospital and Kiambu County Referral Hospital in Kenya between February 2021 and December 2021. This study was part of a multisite clinical study aimed at developing and exploring the use of a paediatric rapid sepsis trigger (PRST) tool. The detailed design and methods of the primary study have been described elsewhere (21).

### Study setting

Mbagathi County Hospital and Kiambu County Referral Hospital are first-level referral hospitals located in Nairobi and Kiambu counties, respectively. The outpatient departments in both hospitals serve approximately 20,000 children annually. Each ED is managed by a qualified nurse who triages children and an additional nurse who administers treatment in the emergency room. In addition, one or two clinical officers (equivalent to physician assistants) provide consultation and decide on the appropriate management of the children. Children triaged as emergency cases by hospital staff are transferred directly to the emergency room, while the rest wait in the queue. Both hospitals admit children to a paediatric ward where provision of care is led by a paediatrician and the clinical team consists of medical officers, nurses, and medical and clinical officer interns. Children who are critically ill and require intensive care are referred to a tertiary hospital for specialized care.

### Population, eligibility, and study procedures

All children aged < 15 years who presented to the ED with an acute illness were eligible to participate. Children scheduled for immunization, elective surgery, wound dressing changes, or clinical review appointments were excluded. Children presenting to the emergency department with an acute illness on weekdays between 8 am and 5 pm were screened for eligibility by study timekeepers and given a sticker showing their arrival time. A systematic sampling method based on 30 minutes time cut-off was adopted to avoid sampling bias. Informed consent was obtained from the caregiver or parent of the first eligible patient at each time cutoff, and assent was obtained from children aged > 13 years. Informed consent and data collection for patients in need of emergency care were deferred, and obtained after the child was stable, to avoid delays.

The study nurses performed clinical examinations and collected data using a password-protected custom-built android application installed on a Samsung Galaxy A8® tablet. Heart rate (HR) and blood oxygen saturation (SpO_2_) were recorded using a Masimo iSpO_2_® pulse oximeter connected to the tablet, and respiratory rate was measured using a version of RRate (22) built directly into the Android data collection application. The children were then reviewed by a qualified clinician who independently decided on the appropriate management. The study nurses recorded hospital outcomes including patient disposition from the hospital records and uploaded the data to a secure REDCap database (23) hosted on the KEMRI Wellcome Trust Research Programme (KWTRP) server. Children who were sent home from the ED on the day of enrolment and those who were admitted were followed up via a telephone call seven days after the initial visit or after discharge from the hospital to ascertain the outcome.

### Triage systems ETAT

This triage system uses clinical signs to assign a triage category depending on the level of illness severity. Frontline health workers identify emergency signs using an “ABCD” method, where A and B symbolize airway and breathing problems, C represents circulation, convulsions, and coma, while D denotes severe dehydration. Children presenting to the hospital with life-threatening problems that require immediate life-saving treatment are assigned to the emergency category, while children requiring urgent review by the health worker (from a set of clinical signs) are assigned a priority category. All other children are classified as non-urgent and can safely wait in the queue (Table 1).

**Table 1:** Clinical signs and categories of the *ETAT* triage system and how they were mapped to our dataset.

| Presenting clinical signs used in ETAT triage | Presenting clinical signs available in the study data | Intervention |
| --- | --- | --- |
| **Emergency** |  | Requires immediate treatment |
| Obstructed/absent breathing | Not available in the baseline dataset |  |
| Central cyanosis | Cyanosis |  |
| Severe respiratory distress | O<sub>2</sub> saturation < 90% OR cyanosis OR grunting OR stridor. |  |
| **Circulation**<br>Capillary refill >3 seconds AND<br>Weak and fast (or absent) pulse AND<br>Cool skin | **Circulation**<br>Capillary refill >3 seconds AND (Weak central pulse OR Weak radial pulse) AND<br>Cool skin |  |
| Convulsions | Convulsions (now) |  |
| Coma | Not alert (based on AVPU scale) |  |
| **Severe dehydration**<br>Diarrhoea plus any two positive signs<br>(Lethargy, sunken eyes, unable to drink or drinks poorly, slow skin pinch) | **Severe dehydration**<br>Diarrhoea plus any two positive signs<br>(sunken eyes, can't sit or drink, slow skin pinch) |  |
| **Priority** |  | Requires prompt assessment |
| Tiny infant (age < 2 months) | Tiny infant (age < 2 months) |  |
| Temperature ≥37.5°C | Temperature ≥37.5°C |  |
| Trauma | Trauma |  |
| Severe pallor | Pallor |  |
| Severe pain | Severe pain |  |
| Poisoning | Poisoning |  |
| Respiratory distress | Chest indrawing<br>OR<br>fast breathing<br>(age < 2 months = RR > 60 breaths per minute<br>age 2-11 months = RR > 50 breaths per minute<br>age ≥ 12 months = RR > 40 breaths per minute) |  |
| Urgent referral | Urgent referral |  |
| Restless, continuously irritable or lethargic | Irritable |  |
| Malnutrition - visible severe wasting | Visible severe wasting |  |
| Oedema of both feet or face | Oedema |  |
| Burns | Burns |  |
| **Non-urgent** |  | Waits in the queue |
| A child without any of the above signs. | A child without any of the above signs |  |

### Smart Triage

This logistic regression model incorporates nine predictors (five continuous and four categorical variables) implemented in a digital device (24). The predictors are age, temperature, heart rate, transformed oxygen saturation, mid-upper arm circumference (MUAC), difficulty breathing, pallor, oedema, and parental concern. The model uses low-risk and high-risk thresholds to categorize children into three triage categories (emergency, priority, and non-urgent). The original model has a low-risk threshold of 8% and high-risk threshold of 40%. This model was previously recalibrated using data from Mbagathi County Hospital which resulted in new thresholds of 2.6% and 13% for low-risk and high-risk thresholds, respectively (manuscript under review). These thresholds were selected to ensure that the model had sensitivity > 80% for identifying high-risk patients. Furthermore, independent emergency and priority triggers (Table 2) were included in a mobile application for the *Smart Triage model* to allow for its safe clinical implementation. The *recalibrated model with independent triggers* is currently being implemented and evaluated at the Mbagathi County Hospital in Kenya.

**Table 2:** Independent emergency and priority triggers included in the Smart Triage.

| Emergency triggers | Priority triggers |
| --- | --- |
| • Unresponsive<br>• Convulsion<br>• Shock (Cool hands with Capillary refill > 3 sec or Weak and fast pulse)<br>• Major trauma<br>• Severe pain<br>• Not breathing<br>• Obstructed breathing<br>• Central cyanosis<br>• Dehydration (At least 2 of sunken eyes, skin pinch taking longer than 2 seconds, or lethargy)<br>• Heart Rate (HR < 45 bpm)<br>• Oxygen Saturation (SpO2 < 90%) | • Trauma or other injury<br>• Burn<br>• Poisoning<br>• Urgent referral<br>• Difficulty breathing (such as Not eating or drinking due to respiratory problems, Chest indrawing, Accessory muscle use, or Head nodding)<br>• Irritable<br>• Respiratory Rate (RR > 60 bpm)<br>• Temperature (Temp > 40°C or Temp < 35°C)<br>• MUAC < 115mm |

### Data analysis

Using data collected from children at Mbagathi County Hospital and Kiambu County Referral Hospital, demographic characteristics were summarized as frequencies, percentages, proportions, medians, and corresponding interquartile ranges (IQR). A classification table was used to compare the distribution of participants into three triage categories according to the ETAT guidelines: the *ST-only model*, the *ST model with independent triggers*, and the *recalibrated ST model with independent triggers*. The change in the classification of the participants into different triage categories was visualized using Sankey diagrams. Smart Triage categories were calculated in Microsoft Excel 2016 (Microsoft, Richmond WA) and then data transferred to R 4.3.0 (R Foundation for Statistical Computing, Vienna, Austria) for final statistical analysis.

### Ethics statement

The study was approved by the Kenya Medical Research Institute (KEMRI) Scientific and Ethics Review Unit (SERU/3958) and Institutional Review Boards at the University of British Columbia in Canada (ID: H19-02398; H20-00484).

## Results

### Participant characteristics

A total of 5920 children (Fig 1) were evaluated for eligibility, of whom 5618 (94.9%) were enrolled in the study. A total 3041 (54.1%) participants were male, and the median age was 20.7 months (IQR 9.0-42.0). Of the enrolled participants, 383 (6.8%) were admitted on the day of enrolment and 11 (0.2%) were readmitted within 48 hours after discharge from the ED (Fig 1). The median length of hospital stay was 6 days (IQR 3-8) (Table 3). Cough was the most common presenting complaint among the participants (1647, 29.4%), while pneumonia was the primary reason for admission (220, 57.4%) (Table 3). Overall, 49 (0.9%) participants died during the study period (Table 3).

**Fig 1:**
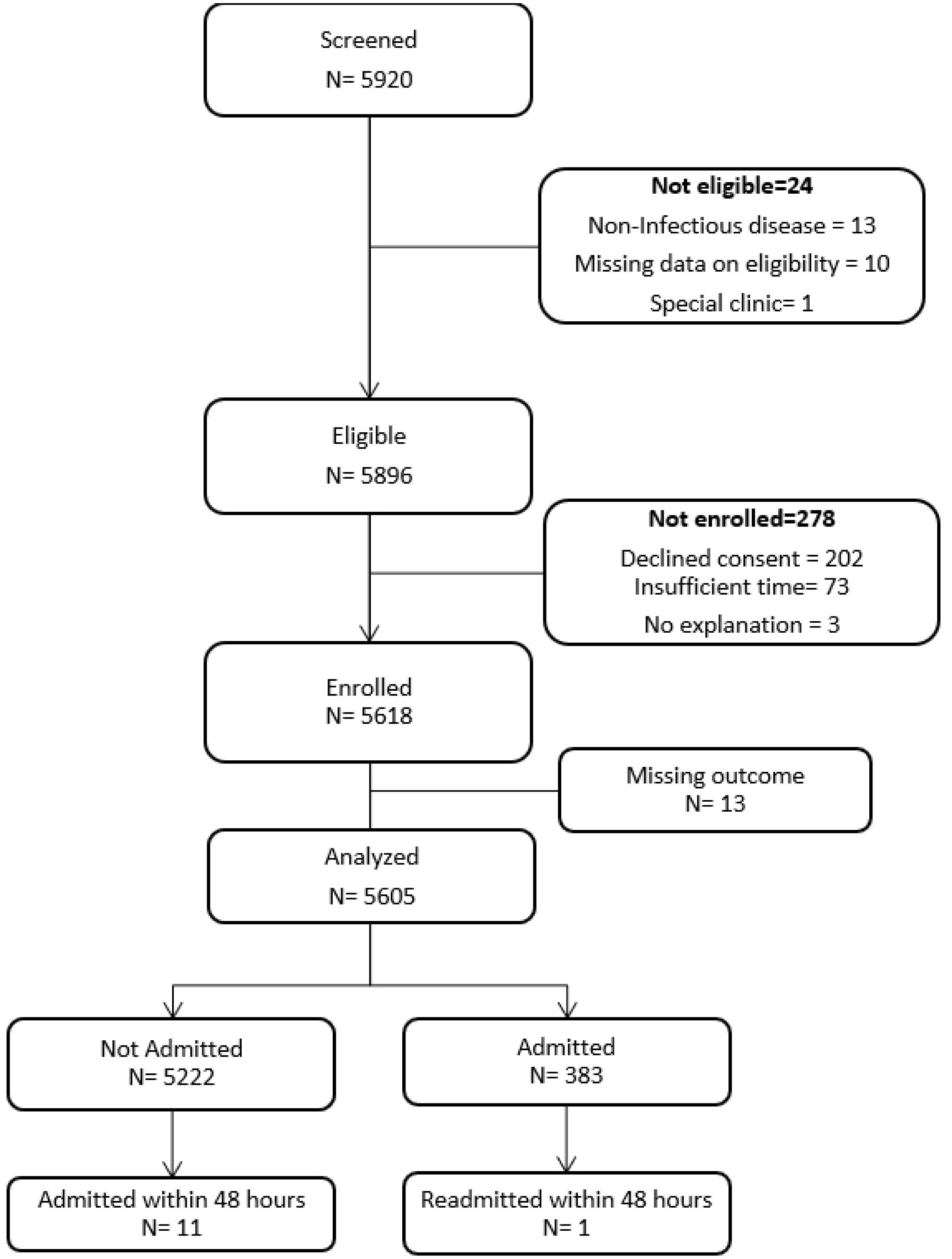
Participant flow diagram.

**Table 3:**
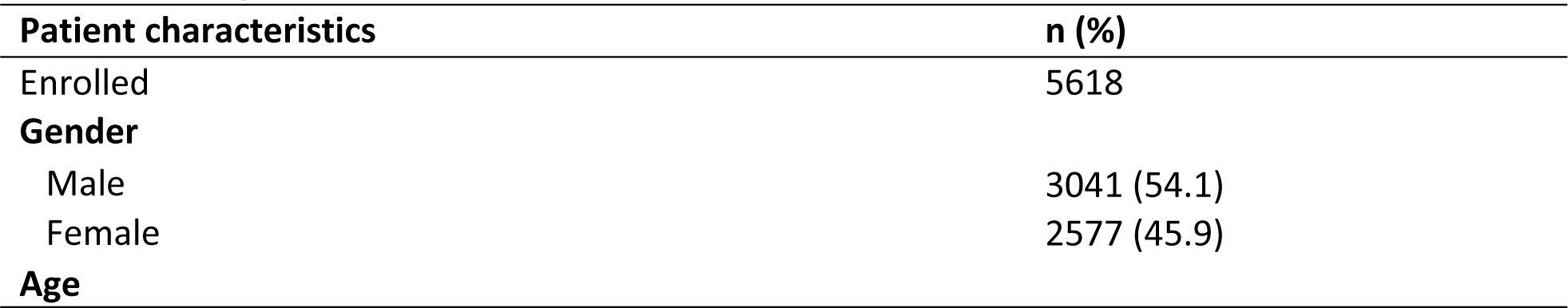
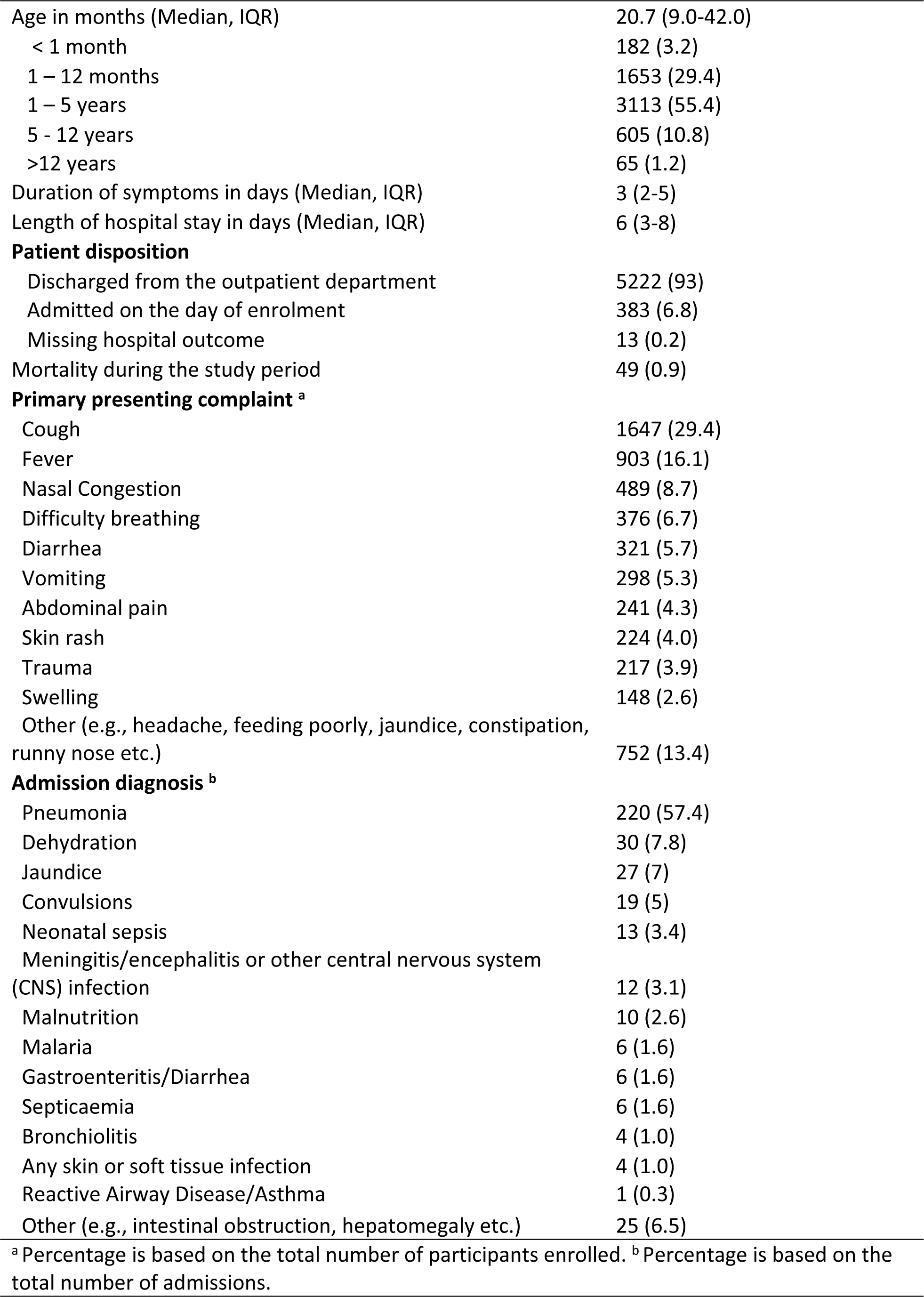
Participant characteristics.

| **Patient characteristics** | **n (%)** |
| --- | --- |
| Enrolled | 5618 |
| **Gender** |  |
| Male | 3041 (54.1) |
| Female | 2577 (45.9) |
| **Age** |  |
| Age in months (Median, IQR) | 20.7 (9.0-42.0) |
| < 1 month | 182 (3.2) |
| 1 – 12 months | 1653 (29.4) |
| 1 – 5 years | 3113 (55.4) |
| 5 - 12 years | 605 (10.8) |
| >12 years | 65 (1.2) |
| Duration of symptoms in days (Median, IQR) | 3 (2-5) |
| Length of hospital stay in days (Median, IQR) | 6 (3-8) |
| **Patient disposition** |  |
| Discharged from the outpatient department | 5222 (93) |
| Admitted on the day of enrolment | 383 (6.8) |
| Missing hospital outcome | 13 (0.2) |
| Mortality during the study period | 49 (0.9) |
| **Primary presenting complaint <sup>a</sup>** |  |
| Cough | 1647 (29.4) |
| Fever | 903 (16.1) |
| Nasal Congestion | 489 (8.7) |
| Difficulty breathing | 376 (6.7) |
| Diarrhea | 321 (5.7) |
| Vomiting | 298 (5.3) |
| Abdominal pain | 241 (4.3) |
| Skin rash | 224 (4.0) |
| Trauma | 217 (3.9) |
| Swelling | 148 (2.6) |
| Other (e.g., headache, feeding poorly, jaundice, constipation, runny nose etc.) | 752 (13.4) |
| **Admission diagnosis <sup>b</sup>** |  |
| Pneumonia | 220 (57.4) |
| Dehydration | 30 (7.8) |
| Jaundice | 27 (7) |
| Convulsions | 19 (5) |
| Neonatal sepsis | 13 (3.4) |
| Meningitis/encephalitis or other central nervous system (CNS) infection | 12 (3.1) |
| Malnutrition | 10 (2.6) |
| Malaria | 6 (1.6) |
| Gastroenteritis/Diarrhea | 6 (1.6) |
| Septicaemia | 6 (1.6) |
| Bronchiolitis | 4 (1.0) |
| Any skin or soft tissue infection | 4 (1.0) |
| Reactive Airway Disease/Asthma | 1 (0.3) |
| Other (e.g., intestinal obstruction, hepatomegaly etc.) | 25 (6.5) |
| <sup>a</sup> Percentage is based on the total number of participants enrolled. <sup>b</sup> Percentage is based on the total number of admissions. |  |

### Triage assignment

Of the total number of participants enrolled, 513 (9.2%), 790 (14.1%), 1163 (20.8%), and 1161 (20.7%) were identified as emergency cases by the *ETAT*, *ST-only* model, S*T model with independent triggers*, and *recalibrated ST model with independent triggers*, respectively (Table 4). *ETAT* categorized the majority of children, 3089 (55.1%), into the non-urgent triage category, unlike the *ST-only* model, *ST model with independent triggers*, and *recalibrated ST model with independent triggers*, which assigned 2442 (43.6%), 2097 (37.4%) and 2617 (46.7%) children, respectively, into the non-urgent category. The *ST-only* model and the *ST model with independent triggers* had a higher proportion of children classified into the priority category, 2373 (42.3%) and 2345 (41.8%), respectively, compared to *ETAT* and the *recalibrated ST model with independent triggers*, which only classified 2003 (35.7%) and 1827 (32.6%) children into this category, respectively. The emergency category had the highest proportion of children who were admitted in all triage systems. For children who died, both the *ST model with independent triggers* and the *recalibrated ST model with independent triggers* identified 39 (79.6%) as emergency cases, whereas *ETAT* and the *ST-only* model identified 25 (51.0%) as such.

**Table 4:** Distribution of participants and outcomes by triage system.

| Triage system | ETAT |  |  | Smart Triage only model |  |  | Smart Triage model with independent triggers <sup>a</sup> |  |  | Recalibrated Smart Triage model with independent triggers <sup>b</sup> |  |  |
| --- | --- | --- | --- | --- | --- | --- | --- | --- | --- | --- | --- | --- |
|  | Emergency | Priority | Non-urgent | Emergency | Priority | Non-urgent | Emergency | Priority | Non-urgent | Emergency | Priority | Non-urgent |
| Participants n (%) | 513 (9.2) | 2003 (35.7) | 3089 (55.1) | 790 (14.1) | 2373 (42.3) | 2442 (43.6) | 1163 (20.8) | 2345 (41.8) | 2097 (37.4) | 1161 (20.7) | 1827 (32.6) | 2617 (46.7) |
| Admission distribution n (%) | 191(17.7) | 168(8.4) | 36(1.2) | 224(28.4) | 134(5.6) | 37(1.5) | 296(25.5) | 83(3.5) | 16(0.8) | 296(25.5) | 74(4.1) | 25(1.0) |
| Mortality on the day of enrolment n (%) | 4(0.7) | 2(0.1) | 0(0) | 4(0.5) | 2(0.1) | 0(0) | 5(0.4) | 1(0) | 0(0) | 5(0.4) | 1(0) | 0(0) |
| In-hospital mortality n (%) | 13(2.5) | 8(0.4) | 0(0) | 14(1.8) | 5(0.2) | 2(0.1) | 18(1.5) | 3(0.1) | 0(0) | 18(1.6) | 3(0.2) | 0(0) |
| Mortality during follow-up n (%) | 8(1.6) | 12(0.6) | 2(0.1) | 7(0.9) | 14(0.6) | 1(0) | 16(1.4) | 6(0.3) | 0(0) | 16(1.4) | 6(0.3) | 0(0) |

<sup>a</sup> **Original Smart Triage model's thresholds:** Non-urgent $\leq 8\%$ , $8\% < \text{Priority} < 40\%$ , Emergency $\geq 40\%$
<sup>b</sup> **Recalibrated Smart Triage model's thresholds:** Non-urgent $\leq 2.6\%$ , $2.6\% < \text{Priority} < 13\%$ , Emergency $\geq 13\%$

### Change in participant classification

#### Overall

Of the 55.1% participants classified as non-urgent by ETAT, 20.1%, 20.4%, and 12.6% were reclassified as priority by the *ST-only model*, *ST model with independent triggers*, and *recalibrated ST model with independent triggers*, respectively (Fig 2a-c). Of the ETAT priority cases, 475 (8.5%) were classified as emergencies by the *ST-only* model. Interestingly, both *ST models with independent triggers* classified 11.7% of the children who had been categorized as priority by *ETAT* as emergency. In addition, for the *ETAT* priority patients, the *ST-only model* assigned more children (465, 8.3%) to the non-urgent category than the *ST model with independent triggers* and the *recalibrated ST model with independent triggers. ETAT* classified 16 (0.3%) children as non-urgent, while the *ST-only model* assigned them as emergency (Fig 2a). Similarly, *ETAT* identified 54 (1%) children as non-urgent, but *ST models with independent triggers* classified them as emergency (Fig 2b-c).

**Fig 2:**
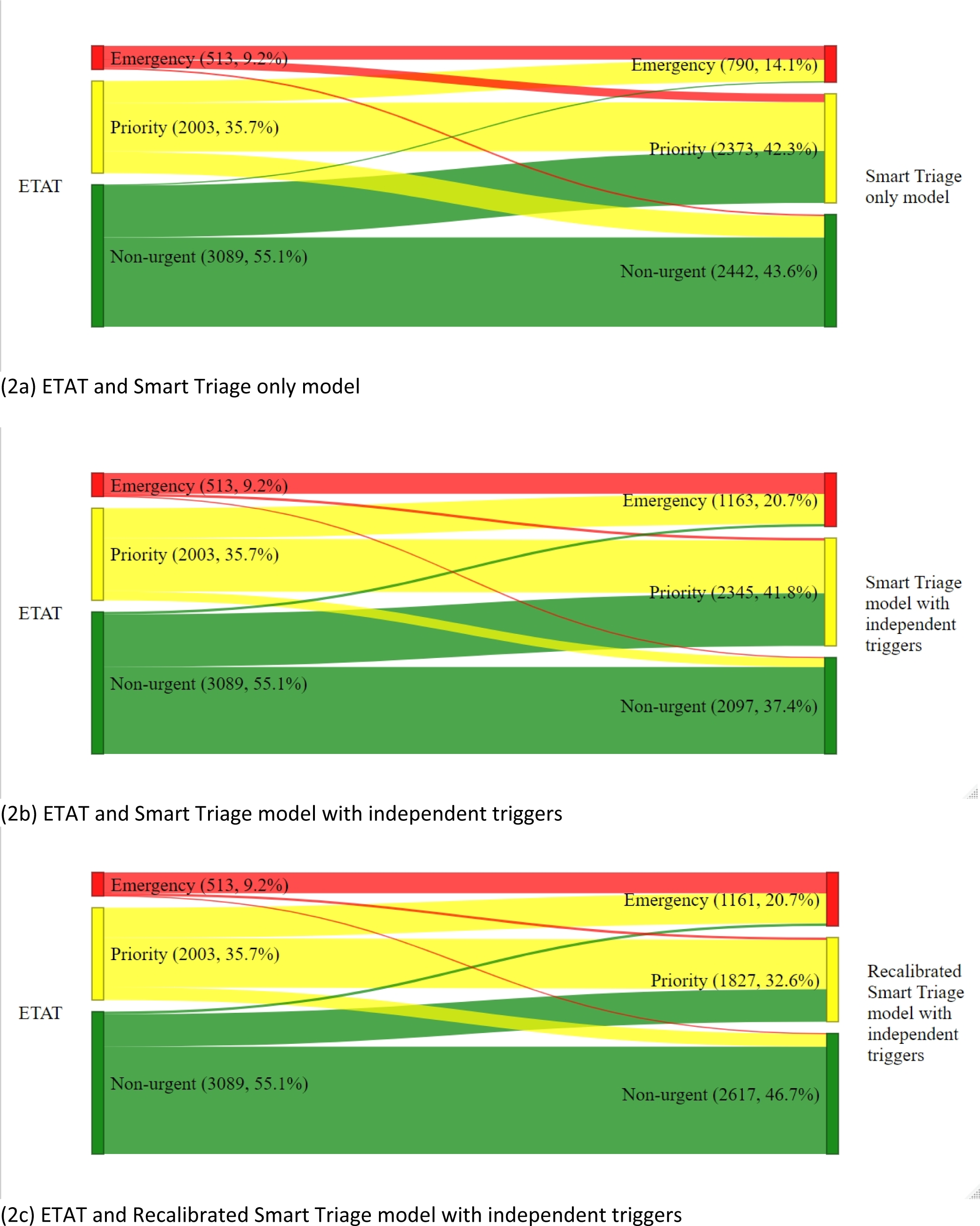
All children. Change in the classification of children presenting to the emergency department by triage systems.

#### Admission

Of *ETAT’s* priority cases, 75/395 (19.0%) and 102/395 (25.8%) of the admitted patients were classified as emergency by *ST-only model* and both *ST models with independent triggers, respectively* (Fig 3a-c). The *ST-only model* categorized 41/395 (10.4%) *ETAT* emergency cases as priority. In contrast, neither the *ST model with independent triggers* nor the *recalibrated ST model with independent triggers* categorized any *ETAT* emergency cases as priority. Overall, the majority of admitted patients were classified into the same triage category using the *ETAT* and *ST models* (Fig 3a-c).

**Fig 3:**
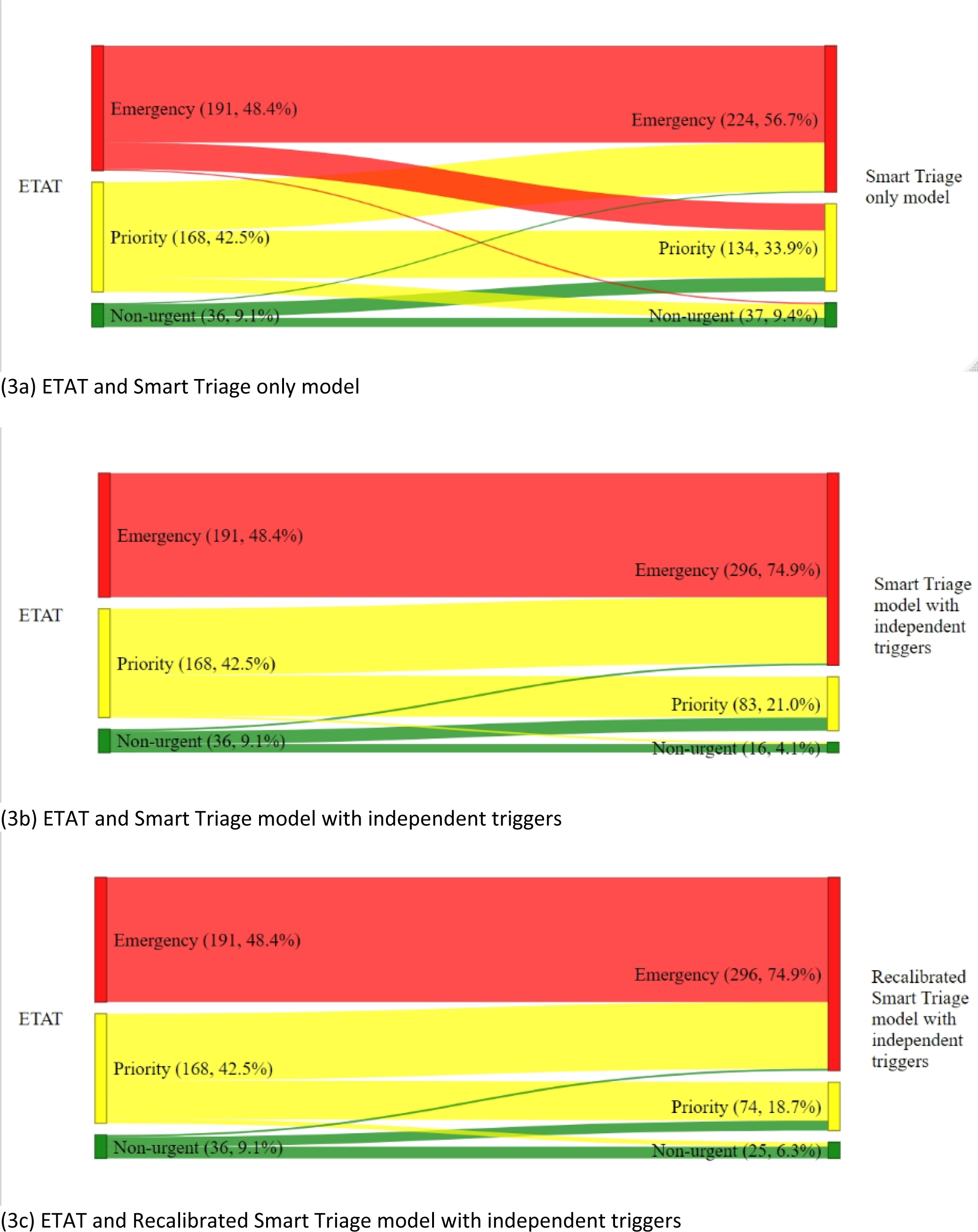
Admitted children. Change in the classification by triage systems for admitted children.

#### Mortality

The *ST models with independent triggers* categorized 34 of the 49 deaths (69.4%) into the same triage categories at *ETAT* (Fig 4). The addition of independent triggers to the Smart Triage model improved the classification of mortality cases by ensuring that all ETAT emergencies were all still emergencies. Of the 49 patients who died, 13 (26.5%) were categorized as priority by *ETAT* and emergency by the *ST models with independent triggers*, whereas 7 (14.3%) were classified as priority by *ETAT* and emergency by the *ST-only model* (Fig 4). The change in the classification of *ETAT* non-urgent patients into other triage categories was similar among *ST models with independent triggers* (Fig 4b-c).

**Fig 4:**
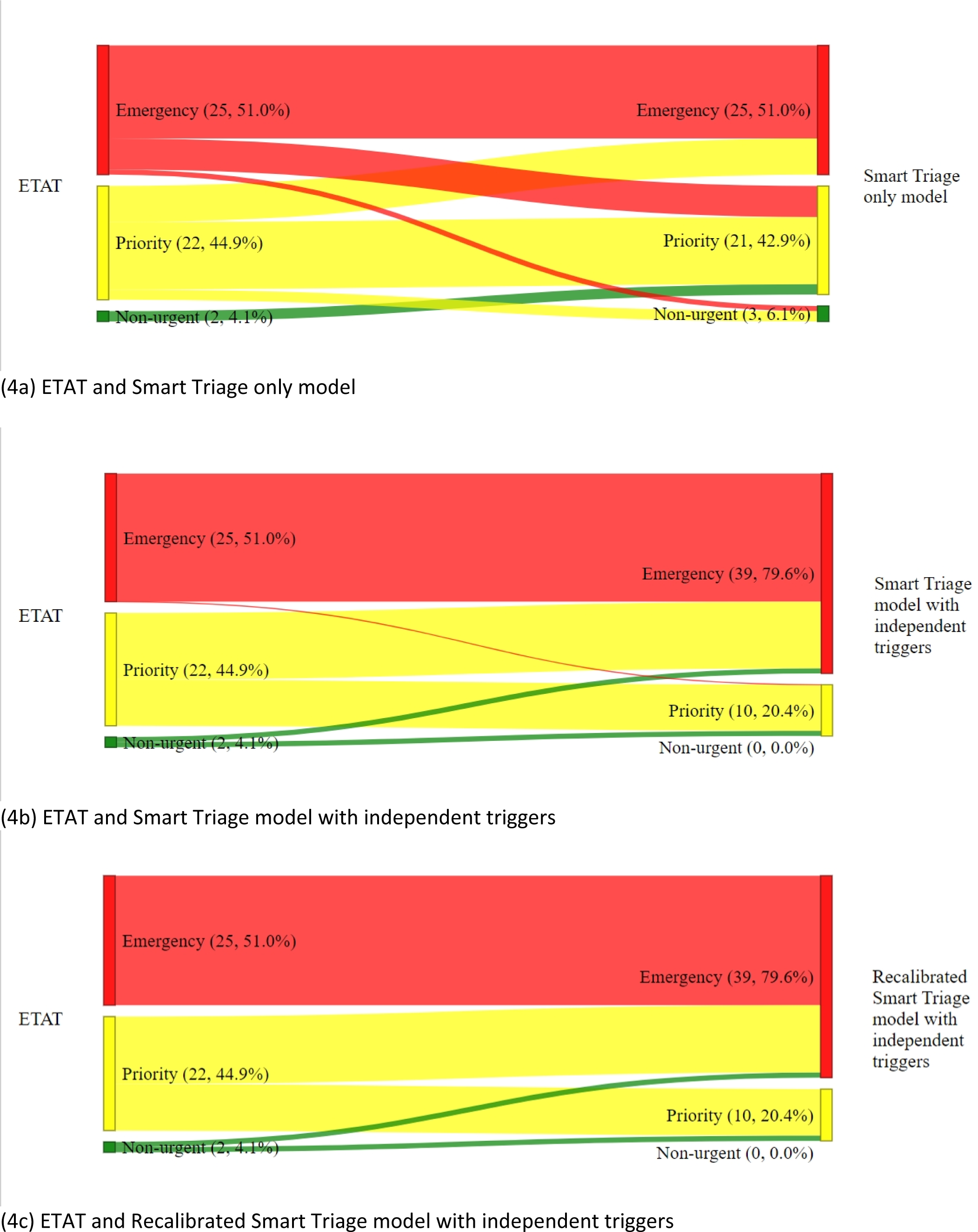
Mortality. The change in the classification by triage system for children who died.

## Discussion

In this study, we compared the *ETAT* guidelines commonly used in Kenyan public hospitals and the newly developed Smart Triage model based on their ability to classify children presenting to the ED into emergency, priority, and non-urgent categories. The major finding based on the selected model thresholds, was a shift in participants classification from non-urgent to priority to emergency when moving from *ETAT* to the *ST-only model*. The magnitude of this shift was larger when independent triggers were added. The emergency category consistently had the highest proportion of children who were admitted or died across all triage systems.

The purpose of paediatric triage is to distinguish children who require urgent intervention by the clinical team from those who can safely wait in the queue when they present to the emergency department. As expected in any triage system, the number of emergency cases should be fewer than priority and non-urgent cases. This was the case in our study, where the proportion of children who were classified as emergency was lower than those assigned priority and non-urgent categories by all triage systems. Assignment of fewer patients in the high acuity category is particularly important to minimize pressure on the already overwhelmed ED. Additionally, the emergency classification expedites clinical review and intervention for children in need of urgent care, thereby improving patient outcomes and ensuring the appropriate use of meager resources.

The Smart Triage models caused a shift in the distribution of children across the three triage categories, with more children being assigned priority categories by the *ST-only model* and the *ST model with independent triggers*. Additionally, all *ST models* classified more children into the emergency category compared to *ETAT*. This was intentionally done with the selection of risk thresholds used to classify patients into these categories, to maximize the models’ sensitivity and specificity, and to reduce misclassification during triage (19). Furthermore, as the Smart Triage algorithm is a continuous probability, the proportion of cases in the emergency and priority groups determined by the algorithm can be adjusted to meet the local context and resources.

Since there is no absolute measure of acuity, admission and mortality have typically been employed in research to assess how effectively a triage system performs (24). Similarly, we used these outcomes as proxies for illness severity. An ideal triage system should categorize children in need of admission or at a high risk of mortality in the high-acuity category. Our study findings showed that *ETAT* and *Smart Triage models* accurately identified critically ill children, with most admissions and deaths assigned to the emergency category. In addition, incorporation of independent triggers improved the Smart Triage model’s ability to classify mortality cases. The *ST model with independent triggers* and the *recalibrated ST model with independent triggers* assigned all mortality cases to either emergency or priority category compared to *ETAT* and *ST-only model* which had some mortality cases assigned to the non-urgent category.

Smart Triage offers a major advantage over *ETAT* in that it can robustly combine a range of continuous and categorical variables in a prediction algorithm that would not be possible for even the most highly trained clinician. The algorithm can also be tuned to optimize the trade-offs clinicians perform in clinical practice. In the current implementation, a high specificity was selected to avoid missing critically ill children, especially those in the early stages of critical illness that have not reached the criteria for priority or emergency status using *ETAT*. This is reflected in the increased number of children in the priority and emergency categories. However, this must be balanced by including too many children in the emergency categories and slowing down access to care for critically ill children with obvious danger signs.

The incorporation of independent emergency and priority triggers into the *Smart Triage models* improved the categorization of admissions and mortality cases, as evidenced by the higher agreement between the *Smart Triage models with independent triggers* and the *ETAT* classification. Most of these triggers are used in *ETAT* to classify patients into emergency and priority categories and can explain the reason for the improvement in the categorization of admission and mortality cases. Therefore, this suggests that the inclusion of independent triggers as an additional criterion in paediatric triage systems that rely on prediction models is essential to avoid missing critically ill children. The prediction algorithm is not necessary in those children with a single symptom or sign that indicates a critically ill child. A similar argument has been proposed when comparing early warning scores to a single extreme observation (25).

### Limitations

This study has some limitations. Enrollment in the study took place on weekdays between 8 am and 5 pm. The distribution of children in different triage categories may differ from that at other times of the day and on weekends. We have used proxies for critical illness such as admission, and in most situations, do not know which triage system was correct. Lastly, the study focused on evaluating the triage systems’ ability to classify children into triage categories and did not assess other important factors, such as resource utilization.

## Conclusion

Smart Triage compares well with *ETAT* and has potential as an efficient system for triaging children and identifying children in need of urgent care, especially when integrated with independent triggers. The Smart Triage algorithm increased the number of children in the priority and emergency groups, but marginally reduced the number of non-urgent children who were admitted or died. The addition of independent triggers to the Smart Triage model further improved the classification of children at high risk of admission and death. Further research is recommended to confirm these findings in other settings, such as primary care, and to compare Smart Triage with other triage systems as well as evaluate its impact on resource utilization.

## Data Availability

The dataset and data dictionary used in this analysis are available upon request from the KEMRI Wellcome Trust Research Programme Data Governance Committee. Applications to access data should be directed to.

## Acknowledgements

We are grateful to all study participants and the administration of Mbagathi County Hospital and Kiambu County Referral Hospital, Charly Huxford, Bella Hwang, Katija Pallot, Metrine Saisi, Elizabeth Kyala, Nancy Githinji, and Caroline Ndolo for their contributions and support during the study period. We highly appreciate the study nurses and clerks: Angela Wanjiru, Anastasia Gathigia, Felix Kimani, Mercy Mutuku, Emmah Kinyanjui, Esther Muthoni, Faith Wairimu, Sidney Kipkorir, Verah Karasi, Victor Achiro, Kevin Bosek, Brian Ochieng and John Mboya for their support in collecting the data.

